# Interest in and preference for long-acting injectable PrEP among men who have sex with men, trans* individuals, and cis-gender heterosexual women: a global systematic review and meta-analysis

**DOI:** 10.1101/2024.09.25.24314401

**Authors:** Haoyi Wang, Johann Kolstee, Alejandro Adriaque Lozano, Liana Aphami, Marco Gaetani, Hanne M.L. Zimmermann, Kai J. Jonas

## Abstract

**Background:** Long-acting injectable HIV pre-exposure prophylaxis (LAI-PrEP) offers potentials for expanding PrEP coverage and improving public health outcomes. This study synthesises global evidence on the prevalence and determinants of interest in and preference for LAI-PrEP among men-who-have-sex-with-men (MSM), trans* individuals, and cis-gender heterosexual women.

**Methods:** We conducted a global systematic review and meta-analysis, building on quantitative studies from a previous review (January 1, 2010, to September 27, 2021) and new studies published in PubMed, Web of Science, and Embase (September 27, 2021, to December 31, 2023). Studies presenting data on LAI-PrEP interest, preference, and their determinants were included. Data were assessed for risk of bias and publication bias. We used a random-effects meta-analysis to pool prevalence of LAI-PrEP interest and preference, and narratively synthesized their determinants across all key populations. This study is registered with PROSPERO, CRD42023488350.

**Findings:** We included 41 articles (18 from the previous systematic review and 23 from the new search). 74% (95%CI 71-78) of MSM showed interest in using LAI-PrEP and 37% (95%CI 29-44) of them preferring LAI-PrEP over other HIV prevention methods. The prevalence of interest and preference were even higher among current oral PrEP users (77% (95%CI 70-84), and 43% (95%CI 28-58), respectively). Interest in LAI-PrEP among trans* individuals was similarly high at 72% (95%CI 67-78), with one study reported that 57% of transgender women preferred for LAI-PrEP. Cis-gender heterosexual women also showed a high preference for LAI-PrEP (55%, 95%CI 40-70)). Overall, people who have more resources and who are already aware and using oral PrEP are likely more interested in LAI-PrEP when it becomes available.

**Interpretation:** Continued research is essential to effectively deploy LAI-PrEP and address HIV prevention gaps. As more countries adopt LAI-PrEP, understanding its impact and reaching underserved populations will be critical to maximising public health benefits.

**Funding:** None.

## Introduction

The availability of advanced HIV biomedical prevention tools is significantly accelerating efforts to end the HIV epidemic.^1^ These tools are particularly crucial for key populations at elevated risk of HIV acquisition,^1,2^ such as men who have sex with men (MSM), trans* individuals and cis-gender heterosexual women.

One of the examples is oral HIV pre-exposure prophylaxis (PrEP).^2^ Daily and on-demand oral PrEP has proven to be highly effective in preventing HIV, and is widely accepted among key populations.^3,4^ By 2023, approximately 3.5 million individuals globally were benefiting from oral PrEP.^1^ However, its uptake varies significantly across regions, with the majority of users in eastern and southern Africa, while other areas are gradually increasing their adoption.^5–7^ Yet, the uptake is still far from reaching the United Nations 2025 targets of on PrEP globally.^1,8^ Additionally, the real-world effectiveness of PrEP is closely associated with its adherence,^9^ which remains a challenge globally.^10,11^ Suboptimal adherence not only reduces the effectiveness of PrEP on the individual level, but also limits its potential public health impact,^12^ creating a significant barrier to HIV prevention efforts.^13^

To address these challenges, novel PrEP modalities, particularly long-acting injectable PrEP (LAI-PrEP), have been developed to alleviate adherence issues and the burden of daily pill-taking. LAI-PrEP has demonstrated superiority over oral PrEP regimens in preventing HIV infection among all the key populations.^14,15^ One such LAI-PrEP option, long-acting injectable cabotegravir, was approved for use as HIV PrEP in the United States (U.S.) and Europe.^16,17^ Additionally, other LAI-PrEP modalities, such as lenacapavir, are under investigation and have demonstrated high efficacy.^18^ Therefore, LAI-PrEP is expected to enhance the public health impact of PrEP among those current users with suboptimal use patterns, and increasing population coverage by attracting individuals who were PrEP-naïve or those who previously discontinued using oral PrEP due to adherence challenges or pill-burdens.^19^

It is therefore crucial to understand the interest in and preference for LAI-PrEP regimens over other HIV prevention methods among key populations to explore their potential for broader global implementation and higher PrEP coverage. A previous systematic review reported a generally high level of interest and preference for LAI-PrEP within these populations.^20^ The review also highlighted significant variations both within and across these key groups. Despite this promising interest, the current evidence remains largely qualitative, only included studies up to September 2021 when there was no authorised LAI-PrEP regimen globally, and mostly based on studies conducted in a U.S. context. Given the increasing interest in long-acting technologies in HIV prevention globally,^21^ there is a clear need for further meta-analytical studies that aim more globally to provide up-to-date robust evidence.

To inform and support future LAI-PrEP research and implementation efforts, it is essential to identify the facilitators and barriers influencing interest and preference for LAI-PrEP. Additionally, it is important to explore whether these determinants differ from those driving interest in and uptake of oral PrEP, as summarised in previous systematic reviews that focused on oral PrEP.^22,23^ A deeper understanding of these variations, particularly those shown to be statistically significant on a global scale, will enable future research to build more effectively on existing findings, leading to more efficient studies and more precise public health strategies. However, to date, no previous systematic reviews have provided these critical insights on LAI-PrEP.

Taken together, there is a clear need for a comprehensive and robust meta-analytical summary of current interest and preference for LAI-PrEP. Additionally, systematically synthesising the determinants of LAI-PrEP interest and preference among key populations is crucial. Therefore, we present an updated global systematic review and meta-analysis, aimed at providing a detailed meta-analytical summary of LAI-PrEP interests and preferences among MSM, trans* individuals, and cis-gender heterosexual women, along with their determinants on a global scale.

## Methods

### Selection criteria and search strategy

This systematic review and meta-analysis is reported following the Preferred Reporting Items for Systematic Reviews and Meta-Analyses (PRISMA) Statement,^24^ and is pre-registered (PROSPERO, ref. CRD42023488350).

For this updated systematic review and meta-analysis, all original studies were eligible for inclusion if they were empirical studies reporting on quantitative or mixed-method analytic findings (quantitative parts) on the proportion or determinants of the interest or preference of long-acting injectable PrEP among MSM, trans* individuals, and cis-gender heterosexual women. Original qualitative studies, opinion pieces, letters to the editors, and other systematic reviews/meta-analyses were not considered eligible for inclusion. To be eligible to be included in our meta-analysis, studies must report the total number of participants, the total number or proportion of participants who were interested in or preferred to use LAI-PrEP. Definitions of LAI-PrEP interest and preference were recorded as a note.

We employed two search methods for this updated systematic review and meta-analysis. Firstly, we built upon the findings from a previously published systematic review of the values and preferences regarding the use of injectable pre-exposure prophylaxis to prevent HIV acquisition.^20^ That review systematically searched for the same endpoints as this updated systematic review and meta-analysis and included articles published between 1 January 2010 and 27 September 2021.^20^ We reviewed all the studies included in that systematic review (n=62). We only included quantitative and mixed-methods studies for our determinant synthesis and meta-analysis. Secondly, we further sought to include articles published between 27 September 2021 and 31 December 2023, by searching PubMed, Web of Science, and Embase, using the combined terms by the population, intervention and outcome (PIO) framework, which included three main constructs: MSM/women/trans*/current oral PrEP users AND injectable modalities/PrEP/prevention AND HIV.

### Data Extraction and Quality Assessment

Data extraction and quality appraisal were performed by AAL, LA, and MG, and verified by HW. The Newcastle-Ottawa Scale for nonrandomised studies was used to assess the methodological quality of the included studies with a cohort study design.^25^

### Meta-analysis

We first extracted the total number of participants for all papers that contained empirical evidence on the number/proportion of the interest or preference of LAI-PrEP. We then extracted the absolute number of participants who reported interest or preference for LAI-PrEP separately by the study population. In terms of multi-centre studies, which reported independent country-specific data, we extracted the data by specific countries instead of treating the multi-centre data homogenously. In terms of studies, which reported independent data on specific sub-populations or intersectional populations, such as MSM and MSM oral PrEP-users, we first extracted the data on the general population, and we then extracted the specific data focus on the sub-populations.

For all meta-analyses, we used a random-effects model and the DerSimonian-Laird method to estimate the model on the proportion. The DerSimonian-Laird *Q* test and *I*^2^ values were used to assess heterogeneity, with low, moderate, and high heterogeneity corresponding to *I*^2^ values of 25%, 50%, and 75%. In addition, heterogeneity τ was assessed in this study. Publication bias was assessed by inspecting funnel plots using a rank correction test. The Statistical analysis was carried out using R (version 4.3.2).

### Determinant systematic synthesis

For all papers that contained empirical evidence on the determinants of LAI-PrEP interest, we first coded the papers based on the study population. Then, we extracted all the determinants/factors the original study reported in the univariable (unadjusted) analyses for each study population. We then extracted all the determinants/factors the original study supported with statistically significant evidence based on the adjusted effect size with 95% confidence intervals or p<0.05. Given the heterogeneous measurements of similar determinants/factors from different included studies, we did not further conduct a meta-analysis to pool the effect size of each determinant. Instead, we summarised all the reported investigated determinants of LAI-PrEP interest among each study population using a narrative approach. We summarised the determinants with the directions of the associations found to be statistically significant in the original studies.

## Results

### Research selection and characteristics

Of the 62 studies included in the previous systematic review,^20^ 18 quantitative studies were included in this systematic review and meta-analysis. Our additional search strategy identified 262 studies after removing duplicates. We excluded 218 studies after screening the titles and abstracts. 43 studies remained for full-text screening, and 20 were excluded, leaving 23 studies to be included. As a result, a total of 41 studies were included in this systematic review and meta-analysis. Figure 1 shows the selection procedure in this study.

Of the 41 studies, 12 originated from middle-low-income countries, four were global multicentre studies and 25 originated from the high-income countries, of which 19 were from the US alone, and only three were from Europe. In addition, of these 41 studies, only two were cohort studies, 35 had a cross-sectional design, two had a mixed-method design, and two were observational studies alongside randomised control studies (Supplementary S1). Quality assessments were summarised in Supplementary S2. There was no evidence of a publication bias in this study.

**Figure 1.**
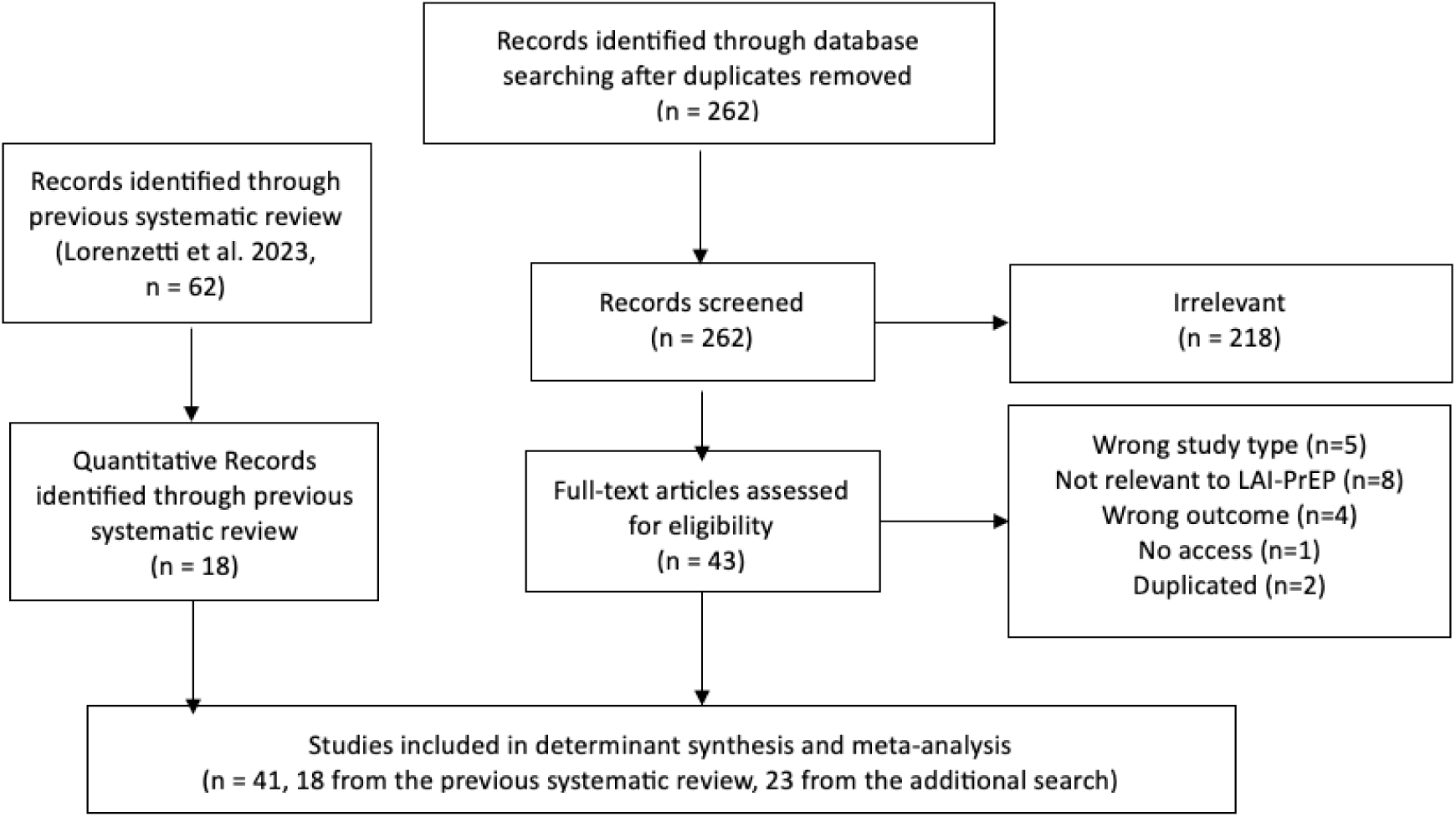
PRISMA Flow diagram of the study selection process.

### Meta-analysis of LAI-PrEP interest and preference

#### LAI-PrEP interest and preference among MSM

Globally, 26 studies reported LAI-PrEP interest among MSM. Based on these studies, a pooled 74% (95%CI0·71-0·78, τ^2^=0·01, I^2^=95·60%) of MSM showed interest in using LAI-PrEP to prevent HIV (Figure 2a). Additionally, 19 studies reported MSM’s preference for LAI-PrEP compared to other HIV prevention methods. A pooled 37% (95%CI0·29-0·44, τ^2^ =0·03, I^2^ =99·40%) of MSM were found to prefer LAI-PrEP to prevent HIV to other HIV prevention methods (Figure 2b).

**Figure 2.**
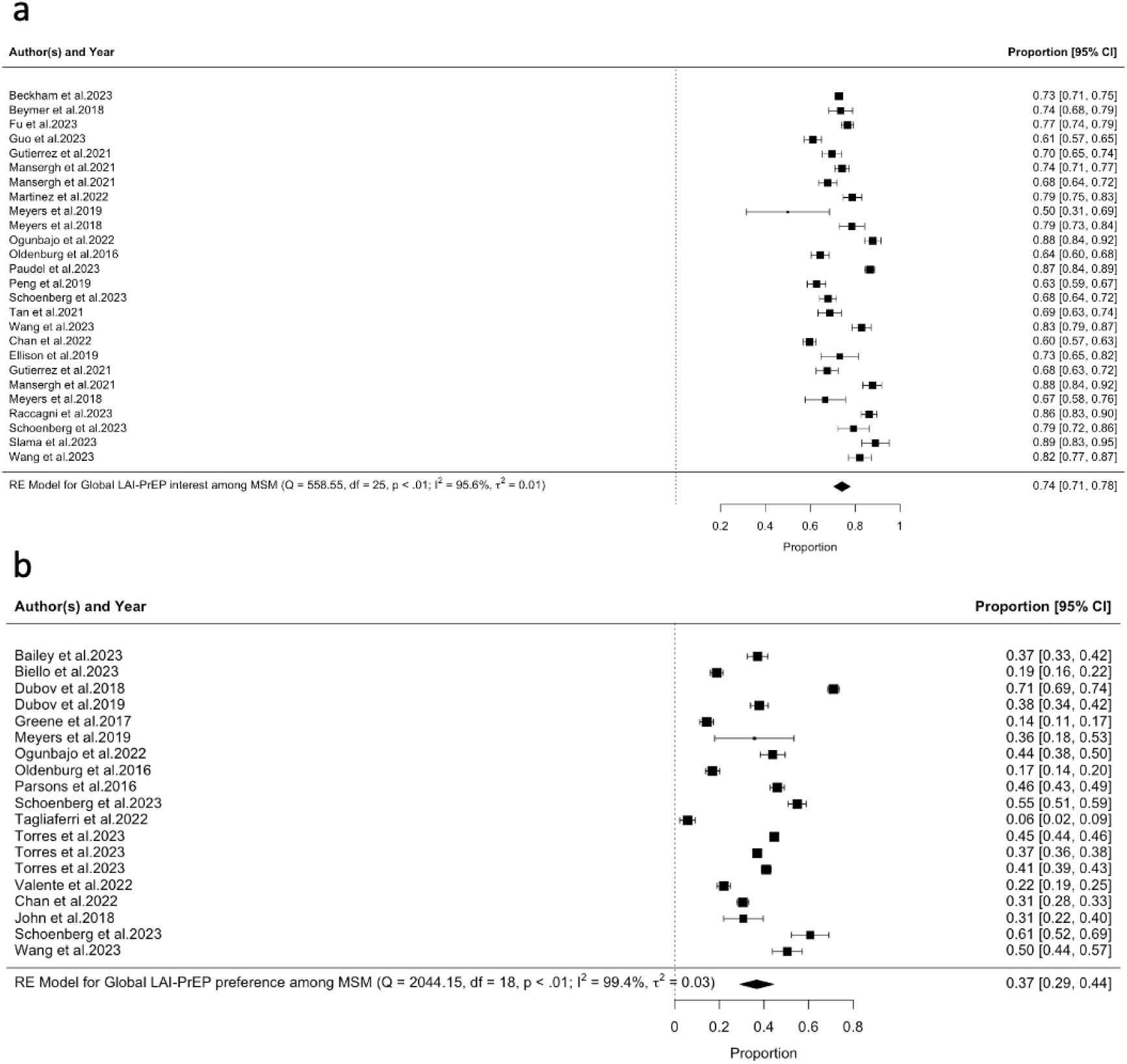
Forest plot of the prevalence of LAI-PrEP a) interest and b) preference among MSM.

#### LAI-PrEP interest and preference among MSM who are current PrEP-users

Globally, nine studies reported LAI-PrEP interest among MSM who are current PrEP-users. Based on these studies, a pooled 77% (95%CI0·70-0·84, τ^2^=0·01, I^2^=94·70%) of them showed interest in using LAI-PrEP to prevent HIV (Figure 3a). Additionally, four studies reported the preference of MSM who are current PrEP-users for LAI-PrEP compared to other HIV prevention methods. A pooled 43% (95%CI0·28-0·58, τ^2^=0·02, I^2^=95·70%) of MSM were found to prefer LAI-PrEP to prevent HIV to other HIV prevention methods (Figure 3b).

**Figure 3.**
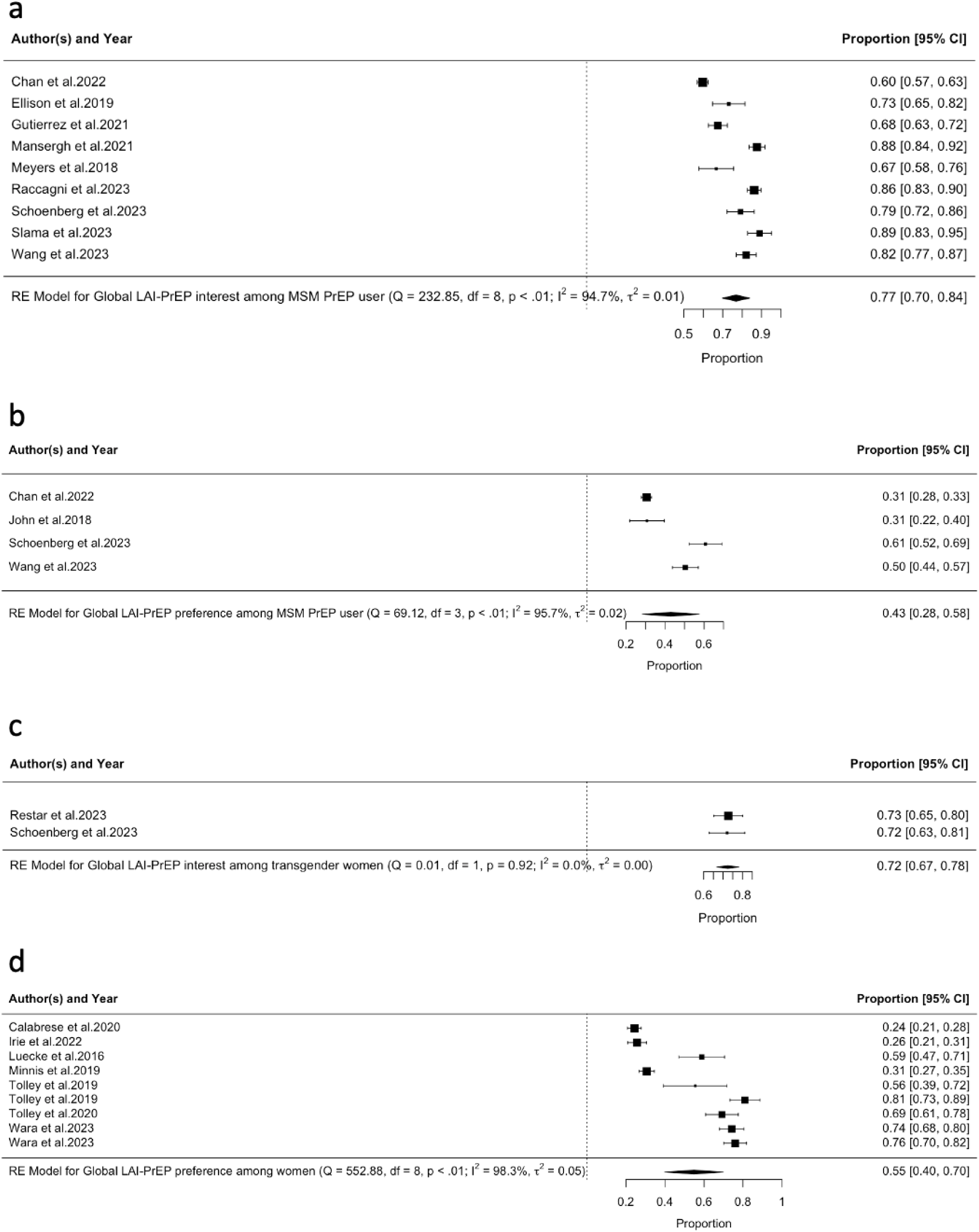
Forest plot of the prevalence of LAI-PrEP a) interest and b) preference among MSM current oral PrEP users, c) LAI-PrEP interest among trans* individuals, and d) LAI-PrEP preference among heterosexual women.

#### LAI-PrEP interest and preference among trans* individuals

Globally, two studies reported LAI-PrEP interest among transgender women. Based on 2 studies, a pooled 72% (95%CI0·67-0·78, τ^2^=0·00, I^2^=0·00%) of transgender women showed interest in using LAI-PrEP to prevent HIV (Figure 3c). In addition, there is only one study that reported LAI-PrEP preference among transgender women. Based on this study, 57% of transgender women would prefer LAI-PrEP to prevent HIV to other HIV prevention methods.^26^ However, no study reported LAI-PrEP interest or preference among transgender men or other gender-diverse populations.

#### LAI-PrEP interest and preference among cis-gender heterosexual women

Globally, based on seven studies, a pooled 55% (95%CI0·40-0·70 τ^2^=0·05, I^2^=98·30%) of cis-gender heterosexual women would prefer LAI-PrEP to prevent HIV to other HIV prevention methods (Figure 3d).

#### Determinants synthesis of LAI-PrEP interest among MSM

Globally, 29 studies reported empirical investigation and evidence on the determinants of LAI-PrEP interest or preference among MSM. More than 75 different sociodemographic, psychosocial, behavioural, and LAI-PrEP-related determinants were investigated in the included studies (Table 1). Of these, 45 were significantly associated with LAI-PrEP interest, with variations among studies.

Among the sociodemographic determinants found to be statistically significant, MSM who were employed, had a primary care provider, higher income, insurance access, or were single, were more likely to express interest in LAI-PrEP. Conversely, MSM who were circumcised or of Muslim faith were less likely to show interest in LAI-PrEP. Evidence was inconsistent regarding the interest in LAI-PrEP among MSM based on their age, education, and ethnicity.

Regarding psychosocial determinants found to be statistically significant, MSM who experienced child sexual abuse, had depression, reported higher levels of fear of HIV, showed interest in psycho-behavioural support services, had higher perceived concern about HIV, struggled with daily pill-taking, were willing to pay for PrEP, and those willing to take daily PrEP, showed higher interest in LAI-PrEP. In contrast, MSM with high concerns about the effectiveness of LAI-PrEP were less likely to be interested.

For behavioural determinants found to be statistically significant, MSM who had engaged in chemsex, condomless anal intercourse (CAI), had ever tested for HIV, experienced monitoring intimate partner violence, had a higher number of sexual partners, were aware of oral PrEP, were current oral PrEP users, early adopters of oral PrEP, were eligible for oral PrEP, sexually active, or used substances, were shown to have higher interest in LAI-PrEP. Conversely, MSM who were older at first intercourse, used condoms, had partners living with HIV, experienced controlling and emotional intimate partner violence, knew someone with HIV, were oral PrEP naive, had optimal adherence to oral PrEP, had a longer history with using oral PrEP, or had engaged in transactional sex, were shown to have lower interest in LAI-PrEP. Evidence regarding former oral PrEP users’ interest in LAI-PrEP was inconsistent.

Regarding LAI-PrEP-related determinants, MSM who found the clinical intervals of LAI-PrEP challenging were significantly less likely to be interested in LAI-PrEP.

**Table 1.**
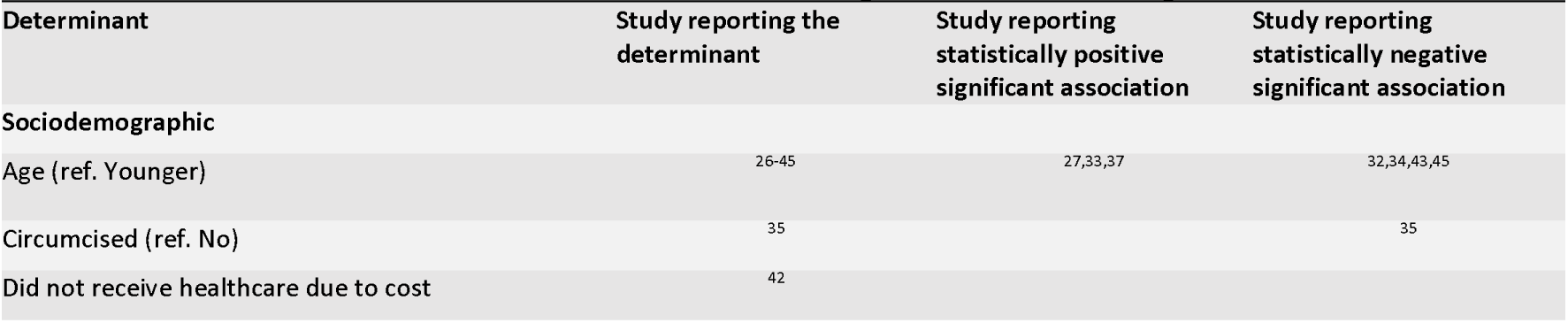
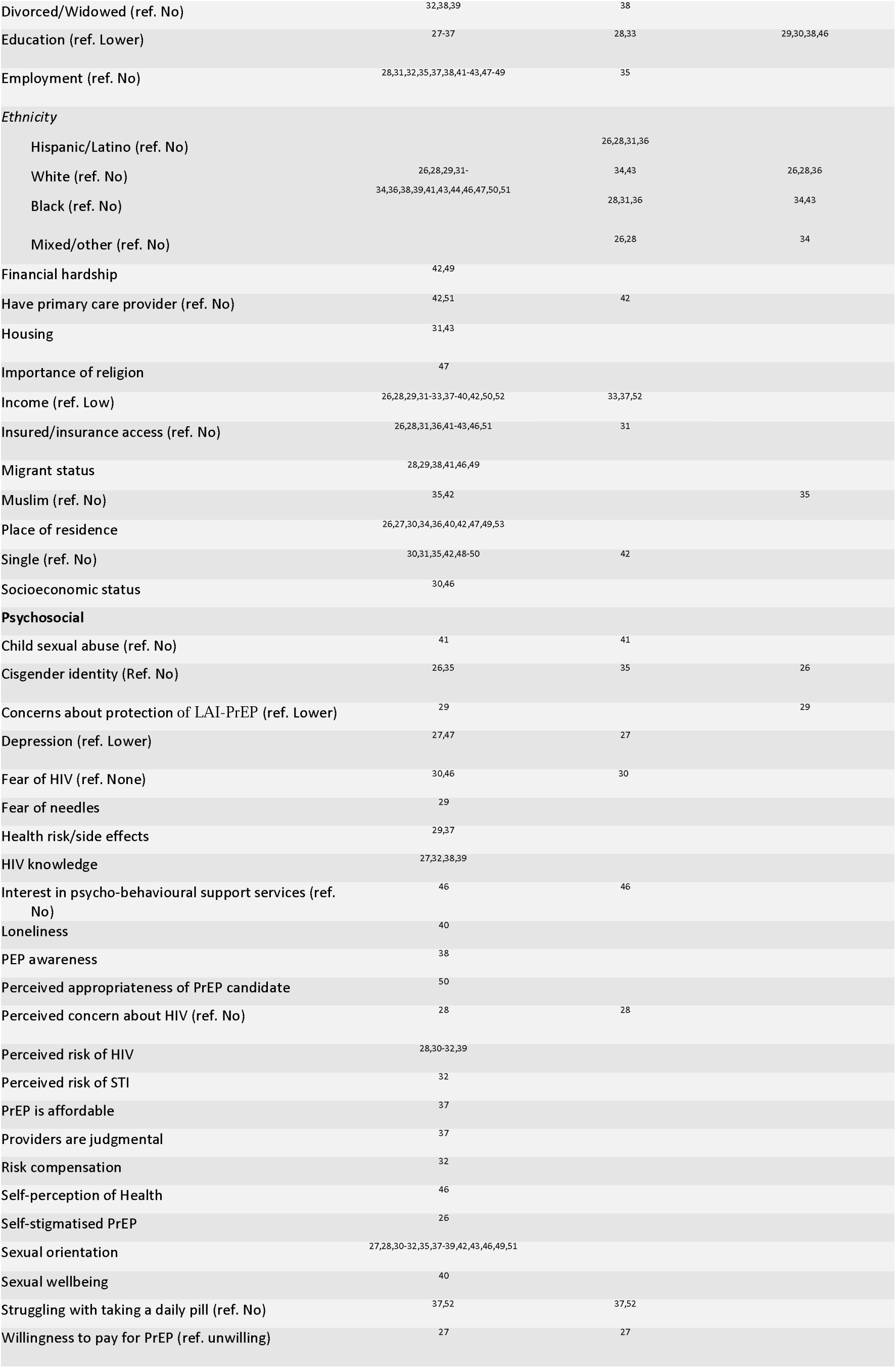
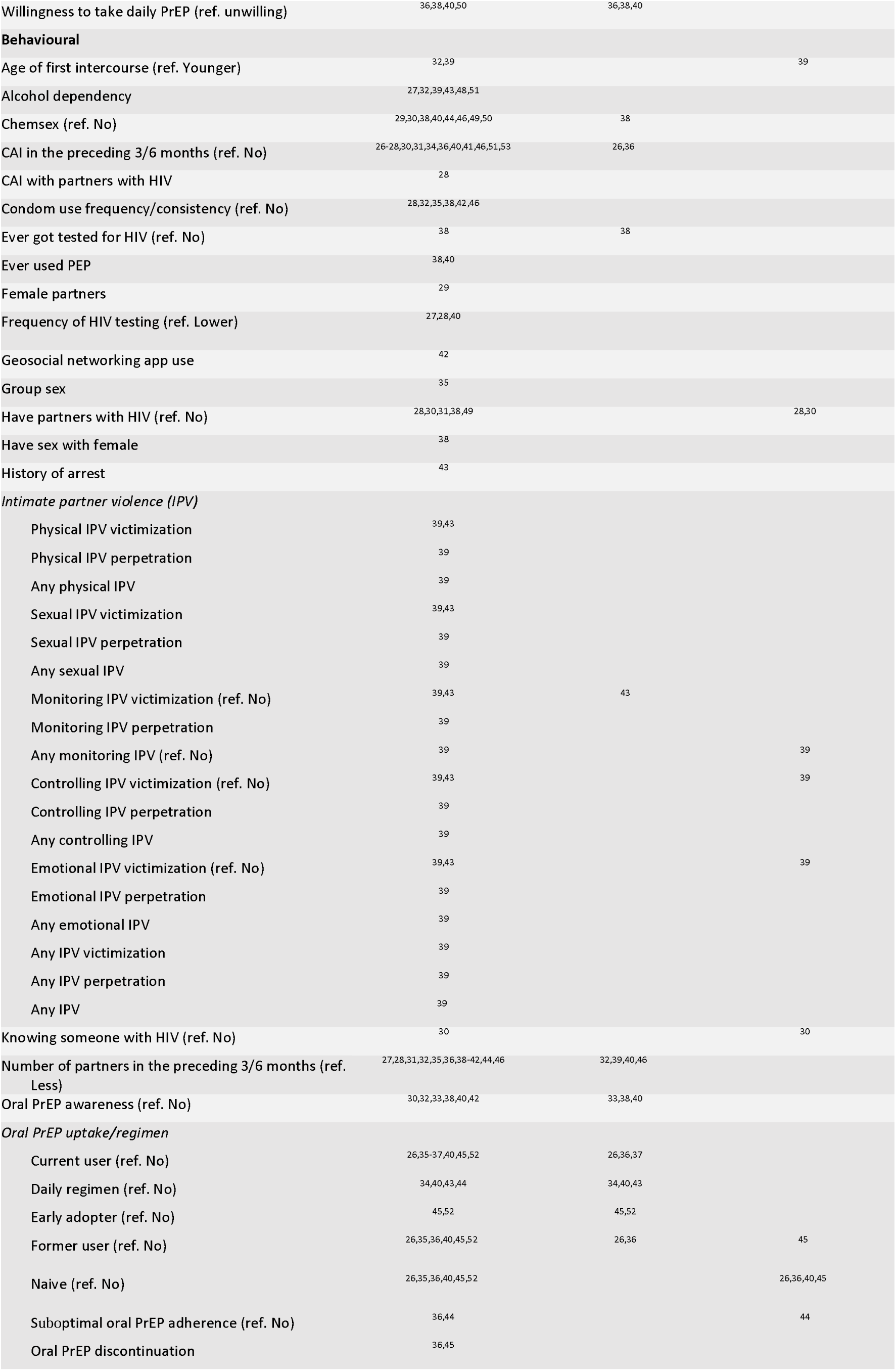
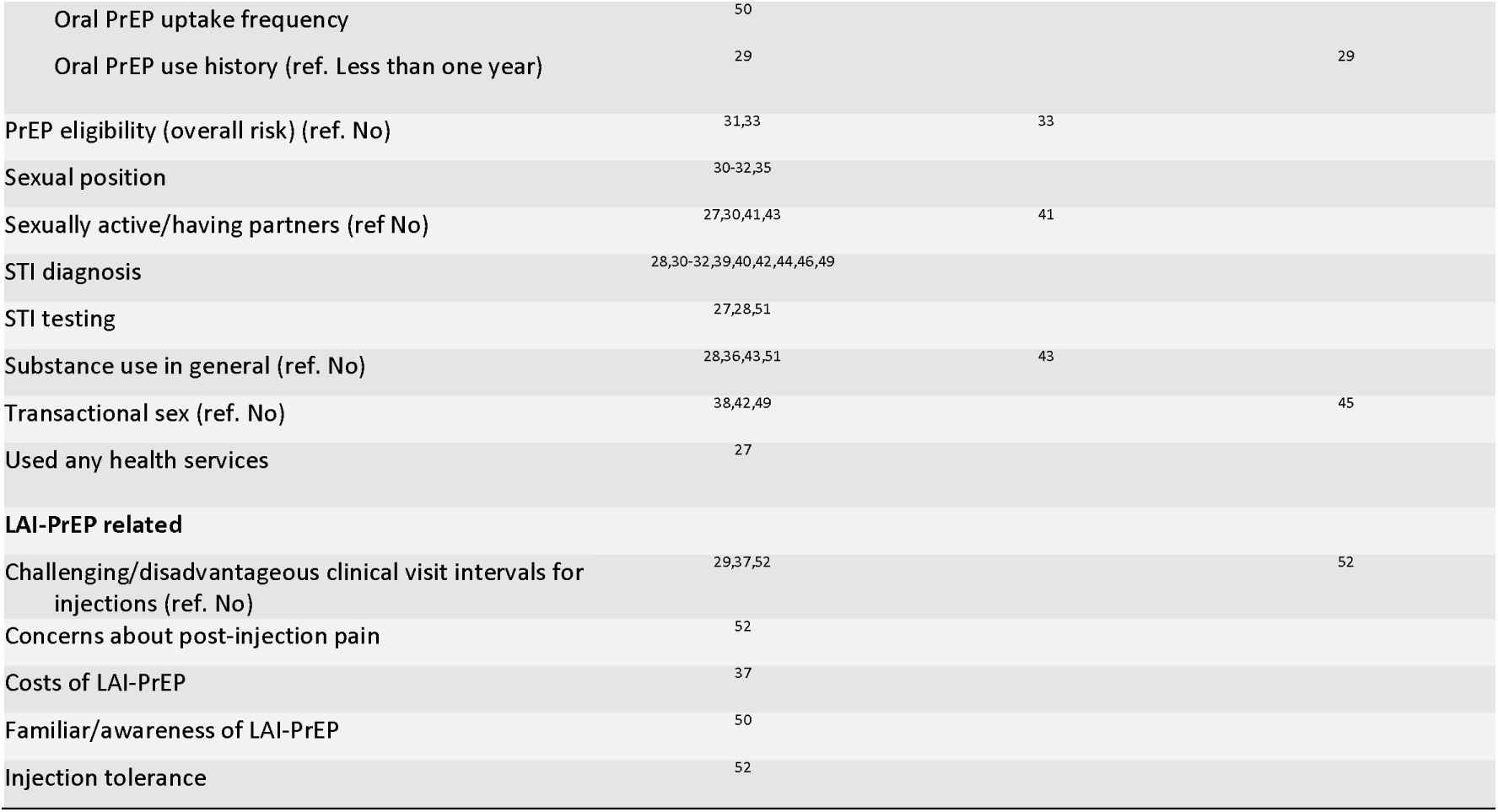
Determinants of LAI-PrEP interest and preference among MSM.

#### Determinants of LAI-PrEP interest among MSM who are current oral PrEP-users

Six studies reported empirical investigation and evidence on the determinants of LAI-PrEP interest among MSM who are current PrEP-users. 35 different determinants were investigated on sociodemographic, psychosocial, behavioural, and LAI-PrEP-related levels, among which 12 were statistically significant (Table 2).

Among the sociodemographic determinants found to be statistically significant, MSM who are current oral PrEP-users who identified as non-Black or Hispanic, had higher education, and were not experiencing financial hardship were more likely to express interest in LAI-PrEP. However, there was no consistent evidence regarding the association between age and interest in LAI-PrEP. Regarding psychosocial determinants, MSM who are current oral PrEP-users who considered daily pill-taking a burden were more likely to express interest in LAI-PrEP. For behavioural determinants, MSM who are current oral PrEP-users who were early adopters of oral PrEP and those with suboptimal adherence to oral PrEP were more likely to express interest in LAI-PrEP, whereas MSM who are current oral PrEP-users with a longer history of oral PrEP use were less likely to be interested in LAI-PrEP. Concerning LAI-PrEP-related determinants, MSM who are current oral PrEP-users who found the clinical intervals of LAI-PrEP challenging, had concerns about its effectiveness, or were uncomfortable with the injection intervals were significantly less likely to be interested in LAI-PrEP.

**Table 2.**
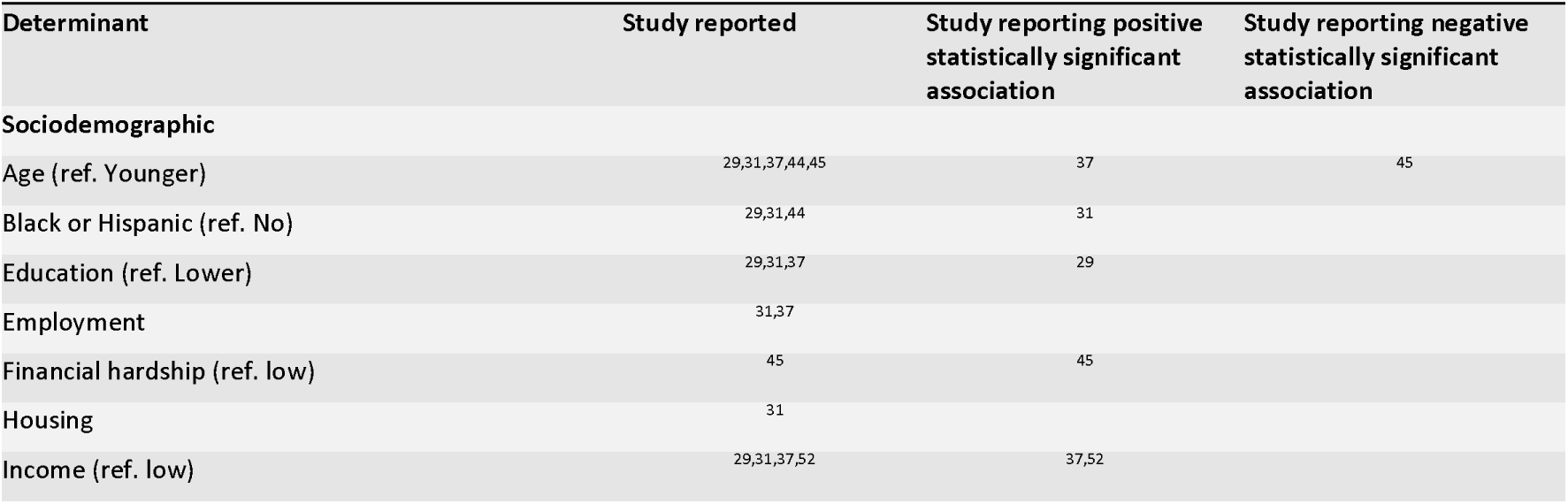
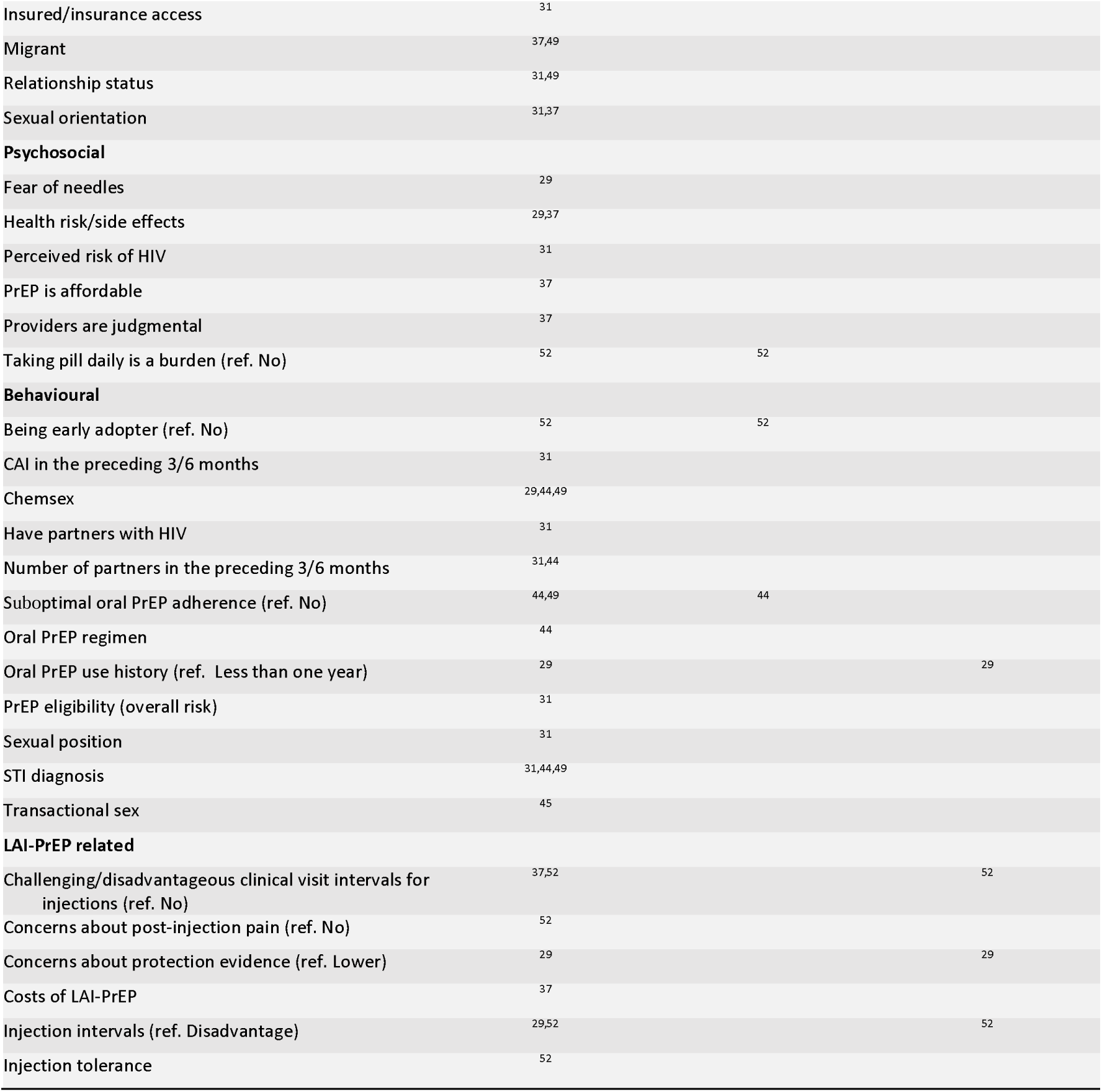
Determinants of LAI-PrEP interest among MSM current oral PrEP users.

#### Determinants of LAI-PrEP interest among cis-gender heterosexual women

Globally, five studies reported empirical investigation and evidence on the determinants of LAI-PrEP interest among cis-gender heterosexual women (Table 3). 29 different determinants were investigated on sociodemographic, psychosocial, behavioural, and LAI-PrEP-related levels, with only eight determinants reported with statistically significant evidence (Table 3).

Among the statistically significant sociodemographic determinants, heterosexual women who received healthcare due to cost and who live outside the middle-and-low income countries such as South Africa or Zimbabwe were more likely to express interest in LAI-PrEP. Conversely, those with higher education and living in the high-income countries such as the US were less likely to show interest. Regarding psychosocial determinants, only cis-gender heterosexual women who experienced PrEP stigma were more likely to have a higher LAI-PrEP interest. For behavioural determinants, cis-gender heterosexual women who reported consistent condom use, those who reported using contraception, and those who had or had a higher number of sexual partners were more likely to express interest in LAI-PrEP. Concerning LAI-PrEP-related determinants, only cis-gender heterosexual women who had higher attributes of LAI-PrEP were likely to express interest in LAI-PrEP.

**Table 3.** Determinants of LAI-PrEP interest among cis-gender heterosexual women.

| Determinant | Study reported | Study reporting positive statistically significant association | Study reporting negative statistically significant association |
| --- | --- | --- | --- |
| **Sociodemographic** |  |  |  |
| Age | 54-56 |  |  |
| Did not received healthcare due to cost (ref. No) | 55 | 55 |  |
| Education (ref. Lower) | 54,56 |  | 54 |
| Employment | 55,56 |  |  |
| Income | 55 |  |  |
| Insurance status | 55 |  |  |
| Marital status | 54 |  |  |
| Place of residence | 54,56,57 |  |  |
| US (ref. No) |  |  | 57 |
| South Africa (ref. No) |  | 54 |  |
| Zimbabwe (ref. No) |  | 54 |  |
| Relationship status | 55 |  |  |
| SES | 54 |  |  |
| **Psychosocial** |  |  |  |
| Altruism | 56 |  |  |
| Comfort discussing PrEP with provider | 55 |  |  |
| Cost to travel to clinic | 57 |  |  |
| Interest in community PrEP delivery | 57 |  |  |
| Perceived concern about HIV | 55,57 |  |  |
| Perceived risk of HIV | 56 |  |  |
| Personal benefits | 57 |  |  |
| Physical experience | 57 |  |  |
| PrEP stigma (ref. No) | 55,56 | 55 |  |
| Time travel to clinic | 56 |  |  |
| **Behavioural** |  |  |  |
| Alcohol dependency | 56 |  |  |
| Condom use (ref. No) | 55-57 | 55 |  |
| Contraception use (ref. No) | 56-58 | 56,58 |  |
| HIV testing | 55 |  |  |
| Number of sexual partners (ref: 0 partner) | 55,56 | 55 |  |
| Oral PrEP history/status | 56 |  |  |
| STI diagnosis | 55 |  |  |
| Transactional sex | 55 |  |  |
| Visited healthcare provider | 55 |  |  |
| **LAI-PrEP related** |  |  |  |
| Product attributes (ref. Low) | 57 | 57 |  |

#### Determinants of LAI-PrEP interest among trans* individuals

Globally, there was only one study reporting empirical investigation of and evidence on the determinants of LAI-PrEP interest among trans* individuals.^59^ The statistically significant determinants identified included discussing HIV services with a healthcare provider and discussing HIV services with sexual partners.

## Discussion

Overall, this analysis, which represents the largest quantitative meta-analytical synthesis on LAI-PrEP interest and preference among key populations to date, found that the majority of MSM, trans* individuals, and cis-gender heterosexual women included in the reviewed studies expressed interest in using LAI-PrEP globally. Additionally, we observed a significant increase in the number of studies on this topic within the past two years compared to the previous decade (23 vs. 18), highlighting the growing importance of generating global evidence on LAI-PrEP interest and preference to inform and enhance HIV prevention strategies worldwide.

Pooled across included studies, interest in using LAI-PrEP was highest among MSM (74%) and even higher among MSM who are current PrEP-users (77%). This high interest in LAI-PrEP may reflect the higher awareness of oral PrEP observed in this group and the normalisation of PrEP use over the past decade among these communities.^2,45,60^ Notably, over a third of MSM (37%) preferred LAI-PrEP to other HIV prevention options, highlighting opportunities to engage this key population further. The higher proportion of current oral PrEP users interested in LAI-PrEP (43%) suggests that many could benefit from switching to this new modality, potentially improving adherence and offering greater convenience. Interest in LAI-PrEP among trans* people was nearly as high as that among MSM (72%), indicating a readiness in this group to adopt this new form of PrEP. The significant preference for LAI-PrEP over other HIV prevention methods among trans* people (57%) further supports its potential to expand HIV prevention coverage in this population. However, with limited studies focusing on trans* individuals, more research is urgently warranted. Encouragingly, cis-gender heterosexual women also showed a high preference for LAI-PrEP (55%), a promising finding given that globally this group is less likely to use PrEP compared to MSM and trans* populations. LAI-PrEP could therefore help address the unmet demand for oral PrEP among this population ^61^. It’s important to note that, despite our efforts to maintain a global focus, nearly half of the studies (19 out of 41) were conducted in the U.S., with very few representing other regions, such as Europe (3 out of 41). This focus is understandable given the availability of Cabotegravir, an LAI-PrEP product, in the U.S. since 2022, whereas it has not yet been officially introduced in many other contexts. This has provided a test case in a jurisdiction where multiple PrEP options are accessible.^16,62^ More research is needed in other high-/middle-income countries, particularly in Europe and East Asia, where LAI-PrEP has been approved by regional authorities. Furthermore, more research is required to understand the interest and opportunities that exist for LAI-PrEP in low-and-middle-income countries in sub-Saharan Africa, Latin America and South/Southeast Asia.

The determinants of interest in LAI-PrEP across studies were diverse but some patterns were observed. Across the studies assessed, MSM were more interested in LAI-PrEP if they came from a more privileged background (e.g., employed, higher income, insured, white), perceived HIV as a threat and believed in the efficacy of PrEP as a preventative tool. In addition, they were more interested in LAI-PrEP, unsurprisingly, if they were sexually adventurous (e.g., higher number of sex partners, practice chemsex) and were already exposed to PrEP either through their own use or awareness of others’ use. Very few studies investigated the determinants of LAI-PrEP interest among cis-gender heterosexual women and trans* people.

The results obtained in this meta-analysis mirror those found in previous studies examining the meanings and values of LAI-PrEP prior to 2021.^20^ These results, based on studies globally over the past three years, demonstrate a growing level of interest in LAI-PrEP among key populations affected by HIV. In particular, MSM and trans* people are aware and knowledgeable about oral PrEP and are ready to pick up new PrEP modalities.

LAI-PrEP provides an incredible opportunity to increase HIV prevention coverage globally, but a targeted promotion of this new modality is required to engage key populations. Understanding who is interested in LAI-PrEP is critical for health systems and the community sectors to understand, so they may most effectively inform people and help them pick up this effective HIV prevention tool. However, further detailed data is needed at the country and regional levels to better determine the geographical differences or variations in LAI-PrEP interests and preferences.

Several limitations exist with this analysis. One key limitation is that in countries where an LAI-PrEP product is not yet available, participants in research studies must provide hypothetical responses regarding their interest and preference for a product they had not yet used. Furthermore, uncertainties around the availability and cost of the product may influence their responses. The rollout of oral PrEP has shown that uptake can be significantly affected by factors such as where the medication can be obtained, who provides it, and how much it cost.^63–65^ Therefore, studies that consider access pathways and the pricing of LAI-PrEP are needed to fully understand their impact on interest and preference. Another limitation is the variation in how LAI-PrEP interest and preference were defined across the included studies. For our meta-analysis, we only included studies that reported absolute numbers of participants interested in or preferring LAI-PrEP, excluding those that measured these factors differently, such as using median scores (e.g., Hsu et al.^40^). This selection could introduce bias into our findings. Finally, another limitation is the lack of differentiation in preferences for different injection intervals of LAI-PrEP, especially as new long-acting injectable modalities are emerging and proving effective.^14,18^ Since most of the included studies focused on a two-monthly LAI-PrEP regimen, our findings may not be generalisable to newer, twice-yearly LAI-PrEP options.

## Conclusion

In conclusion, our analysis found high levels of interest in LAI-PrEP among key populations affected by HIV, though notable differences persist between groups and regions. Overall, people who have more resources and who are already aware and using oral PrEP are likely more interested in LAI-PrEP when it becomes available. Continuing to build the evidence base will be crucial to understanding how to best deploy this new HIV prevention tool. As the HIV epidemic continues to evolve, communities continue to respond to it, and as more countries make LAI-PrEP available, ongoing research will be needed to understand the HIV prevention gaps that remain.

## Data Availability

All data produced in the present study are available upon reasonable request to the authors

## Declaration of interests

### Ethical statement

This systematic review and meta-analysis is reported following the Preferred Reporting Items for Systematic Reviews and Meta-Analyses (PRISMA) Statement, and is pre-registered (PROSPERO, ref. CRD42023488350).

### Authors’ contributions

HW, JK, HMLZ, and KJJ conceptualised this research; HW, AAL, LA and MG collected the data for this research; HW analysed the data; HW, JK and KJJ drafted the manuscript; All authors critically revised the manuscript for intellectual content; All authors read and approved the final version of the manuscript.

## Acknowledgements

None.

