## Supplementary File for "Interest in and preference for long-acting injectable PrEP among men who have sex with men, trans* individuals, and cis-gender heterosexual women: a global systematic review and meta-analysis"

**Table S1. Study characteristics**

| **Author** | **Aim** | **Country/region** | **Study Design** | **Data Collection** | **Study Sample** | **Outcome of Interest** | **Notes** |
| --- | --- | --- | --- | --- | --- | --- | --- |
| Bailey et al. (2022) ^33^ | Evaluate preferences for current and future HIV prevention options and assess socio-demographic and behavioural factors associated with each preference | Kenya | Cross-sectional | 2021 | 464 HIV negative MSM and transgender women | LAI-PrEP preference |  |
| Beckham et al. (2023) ^34^ | Understanding variations in potential user preferences for LA PrEP via latent class analysis | US | Cross-sectional | 2019 | 2,489 MSM | LAI-PrEP interest |  |
| Beymer et al. (2018) ^26^ | Assess the willingness to try alternative PrEP delivery mechanisms among YMSM | US | Cohort study | 2015 | 761 YMSM | LAI-PrEP interest |  |
| Biello et al. (2023) ^49^ | Examine the preferences for features of PrEP products within YMSM participants via conjoint analy- ses and across PrEP product preferences and correlates of these preferences between YMSM participants (via rank- choice selection) | US | Cross-sectional | 2020-2021 | 737 cis-gender YMSM | LAI-PrEP preference | Rank-ordered regression was used, complicating comparison of the results with those of other studies |
| Calabrese et al. (2019) ^56^ | Investigate women’s preferences across 10 PrEP modalities currently available or under study and examine associations between PrEP modality preferences and contraception practices | US | Cross-sectional | 2017 | 563 heterosexual women | LAI-PrEP preference |  |
| Chan et al. (2022) ^35^ | Investigate the interest and most preferred modality of PrEP modalities amidst daily dose, on-demand, long-acting injectable and implants | Australia | Cross-sectional | 2019-2020 | 1477 GBM current PrEP users | LAI-PrEP interest and preference |  |
| Dubov er al. (2018) ^64^ | To elicit MSM preferences in order to inform program development to facilitate successful delivery of PrEP to Ukrainian MSM | Ukraine | Cross-sectional | 2018 | 1184 MSM | LAI-PrEP preference |  |
| Dubov er al. (2019) ^45^ | To elicit MSM stakeholder preferences in order to inform program development aimed at improving uptake of PrEP. | US | Cross-sectional | 2019 | 554 MSM | LAI-PrEP preference |  |
| Ellison et al. (2019) ^29^ | Evaluate the barriers in using and adhering to oral PrEP, preferences for PrEP modalities, sociodemographic characteristics and sexual behaviors related to these preferences, and the reasons behind their interest or disinterest in each PrEP modalities | US | Cross-sectional | 2017 | 108 MSM current PrEP users | LAI-PrEP preference |  |
| Fu et al. (2023) ^36^ | Explore the willingness to use LAI-PrEP and associated influential factors | China | Cross-sectional | 2020-2021 | 969 HIV-negative MSM | LAI-PrEP interest |  |
| Greene et al. (2017) ^58^ | Identify preferences for current and future HIV prevention options and explore reasons for preferences among HIV prevention options including potential new long-acting delivery options | US | Cross-sectional | 2017 | 512 cis-gender gay or bisexual MSM | LAI-PrEP preference | No univariate or multivariate analysis (e.g. regression) testing the role of the variables included in predicting the preference for LAI-PrEP was conducted. |
| Guo et al. (2023) ^37^ | Investigate the prevalence of the intention to use different types of PrEP and explore the associations between IPV experience and willingness to use PrEP | China | Cross-sectional | 2018-2019 | 608 HIV-negative MSM | LAI-PrEP interest |  |
| Guttierrez et al. (2022) ^65^ | To identify population-based, segment-specific preferences for longer-acting and alternative PrEP delivery modalities to guide patient-centered strategies to optimize uptake within military-serving healthcare systems. | US | Cross-sectional | 2020 | 429 militery MSM | LAI-PrEP interest |  |
| Hsu et al. (2023) ^38^ | Determine the preferences for the different forms and dosing intervals of LAI-PrEP that are currently in the development pipeline | Taiwan, China | Cross-sectional | 2021 | 1728 HIV-negative MSM | LAI-PrEP preference |  |
| Irie et al. (2022) ^53^ | Evaluate preferences for different PrEP products, including daily pills, long-acting injectables, vaginal gels, and vaginal rings. Focused on the prevalence and reasons for preferring LAI PrEP to oral PrEP, considering factors like sexual behavior, stigma, and healthcare access | US | Cross-sectional | 2019 | 315 cis-gender black women who have sex with men | LAI-PrEP preference |  |
| John et al. (2018) ^27^ | Assess the awareness and preferences regarding LA-PrEP among GBMSM who are currently using oral PrEP | US | Cross-sectional | 2015-2016 | 104 GBM current PrEP users | LAI-PrEP preference |  |
| Luecke et al. (2016) ^52^ | Investigate the insights of women's preferences and behaviors regarding HIV prevention products to inform future HIV prevention strategies and product development | Multicenter | Mixed-method | 2013-2014 | 68 cis-gender women | LAI-PrEP preference |  |
| Mansergh et al. (2021) ^32^ | Assess self-reported likelihood of using various HIV prevention products, including a LAI form of PrEP and sexual event-based pills, gels and anal suppositories | US | Cross-sectional | 2018 | 782 HIV-negative MSM | LAI-PrEP interest |  |
| Martinez et al. (2022) ^39^ | Investigate what factors are correlated with interest in four biomedical HIV prevention methods (including dissolvable implants, removable implants, rectal douching and injection) among racially diverse, and  look at the demographic characteristics and sexual risk behaviours among a racially diverse sample of MSM | Northeast Corridor region between Philadelphia and Trenton | Cross-sectional | 2022 | 381 MSM | LAI-PrEP interest |  |
| Meyers et al. (2014) ^44^ | Investigate interest in and attitudes towards LAI-PrEP | US | Cohort study | 2013 | 197 HIV-negative YMSM | LAI-PrEP interest |  |
| Meyers et al. (2016) ^50^ | To investigate which factors might help direct a patient-physician shared-decision making process to optimize the choice of biomedical HIV prevention method. | US | Cross-sectional | 2017 | 105 current oral PrEP users | LAI-PrEP interest |  |
| Meyers et al. (2018) ^28^ | Explore factors influencing the selection of biomedical HIV prevention methods, particularly focusing on generating data to distinguish between current oral PrEP users considering a switch to LAI-PrEP, with an emphasis on product-related and psychosocial factors affecting LAI-PrEP implementation | US | Cross-sectional | 2014-2017 | 105 cis-gender MSM who are currently using oral PrEP | LAI-PrEP interest |  |
| Meyers et al. (2018) ^28^ | Investigate interest in and attitudes toward LAI-PrEP and how men only interested in the injectable formulation of PrEP differ from those who would use PrEP in any modality or not at all | China | Cross-sectional | 2013-2014 | 200 HIV-negative MSM | LAI-PrEP interest |  |
| Minnis et al. (2019) ^66^ | To examine youths’ preferences for key attributes of long‐acting Pre‐Exposure Prophylaxis (PrEP), with a focus on characteristics pertinent to product delivery alongside key modifiable product attributes | South Africa | Cross-sectional | 2018-2019 | 401 heterosexual women | LAI-PrEP preference |  |
| Ogunbajo et al. (2022) ^40^ | Investigate the association between demographics, socioeconomic marginalization, and sexual health and willingness to use LAI-PrEP and preferences for other PrEP modalities | Nigeria | Cross-sectional | 2019 | 305 HIV-negative or serostatus unknown SMM | LAI-PrEP interest and preference |  |
| Oldenburg et al. (2016) ^25^ | Assess preferences for daily oral, injectable, and rectal microbicide modalities of PrEP, as well as participants’ view on potential barriers to uptake and adherence | Vietnam | Cross-sectional | 2015 | 548 cis-gender and HIV-negative MSM | LAI-PrEP preference |  |
| Peng et al. (2019) ^30^ | Investigate the level of willingness to use three types of PrEP (daily oral PrEP, on-demand PrEP, and LAI-PrEP) and their associated factors, and the level of intention to adhere to PrEP usage with these three types and the associated factors | China | Cross-sectional | 2018-2019 | 524 HIV-negative or serostatus unknown MSM | LAI-PrEP interest |  |
| Raccagni et al. (2023) ^42^ | Investigate the predisposition in switching to LAI-PrEP with cabotegravir every two months. Assess the factors associated with the switch | Italy | Cross-sectional | 2023 | 377 cis-gender MSM current PrEP users | LAI-PrEP interest |  |
| Restar et al. (2023) ^57^ | To explore PrEP awareness, discussion and interest in taking LAI-PrEP among Filipina transfeminine adults. | Philippines | Cross-sectional | 2022 | 139 transgender women | LAI-PrEP interest |  |
| Schoenberg et al. (2023) ^24^ | Assess the acceptability of LAI-PrEP among SGM people in urban and non-urban areas, and describe differences in willingness to use LAI-PrEP and preference for PrEP modality based on demographics, including race and ethnicity, gender identity, age, socioeconomic characteristics, and sexual health history | US | Cross-sectional | 2022 | 583 HIV-negative or serostatus unknown cis- and trans-gender males and transgender or non-binary females | LAI-PrEP interest and preference |  |
| Slama et al. (2023) ^46^ | To focus on patient perspectives and to understand which individuals, among PWH and PrEP users, would constitute the preferential target for such treatments in terms of expectations but also tolerability, adherence and improvement in quality of life. | France | Cross-sectional | 2023 | 100 MSM | LAI-PrEP interest |  |
| Stephenson et al. (2022) ^41^ | Examine how the recent experience of IPV may shape rankings of PrEP delivery options and explore how different forms of IPV may shape preferences for PrEP modalities | US | RCT | 2018-2019 | 694 cis-gender GBMSM | LAI-PrEP preference |  |
| Tagliaferri et al. (2022) ^67^ | To estimate the preference share for the implant within a competitive context of other PrEP products (including the oral tablet, dissolvable implant, and injection) and evaluate the impact of potential implant attributes. | US | Cross-sectional | 2022 | 175 MSM | LAI-PrEP preference |  |
| Tan et al. (2021) ^68^ | To characterize preferences for existing and forthcoming PrEP modalities among gbMSM in Toronto. | Canada | Cross-sectional | 2016 | 303 MSM | LAI-PrEP interest |  |
| Tolley et al. (2019) ^55^ | Assess the acceptability of HIV prevention products based on user perceptions, perceived prevention needs, and product familiarity | Multicenter | Mixed-method | 2015-2017 | 136 cis-gender women | LAI-PrEP interest and preference |  |
| Tolley et al. (2020) ^69^ | To compared acceptability of product attributes, prevention preferences and future interest in injectable PrEP | US | Cross-sectional | 2020 | 117 heteroseuxal women | LAI-PrEP preference |  |
| Torres et al. (2023) ^31^ | Assess preferences for PrEP modalities | Multicenter | Cross-sectional | 2018 | 19,457 MSM | LAI-PrEP preference |  |
| Valente et al. (2022) ^70^ | To identify latent classes of YMSM based on characteristics of sexual health decision-making and experiences accessing health care; and to examine associations between profiles of YMSM and engagement in health care and HIV prevention, and preferences across PrEP modalities. | US | Cross-sectional | 2020-2021 | 737 heteroseuxal women | LAI-PrEP preference |  |
| Wang et al. (2024) ^43^ | Investigate the intention in using LAI-PrEP by comparing MSM who are PrEP naïve, discontinued oral PrEP, and are early adopters and the majority oral PrEP users, using the diffusion of innovation approach | The Netherlands | Cross-sectional established within a cohort study | 2022 | 309 MSM who are current oral PrEP users | LAI-PrEP interest and preference |  |
| Wara et al. (2023) ^54^ | Assess the preference and perception of long-acting modality | Africa | Observational cohort study / Observational extension cohort of a cluster randomized trial study | 2021-2022 | 394 cis-gender women who are currently using or previously used daily oral PrEP and are currently pregnant or postpartum | LAI-PrEP preference |  |

Note: LAI = long acting injectable; SMM = sexual minority men; MSM = men who have sex with men; SGM = sexual and gender minority; GBMSM = gay, bisexual and other men who have sex with men; YMSM = young men who have sex with men; IPV = intimate partner violence.

**Table S2. Newcastle-Ottawa quality assessment of non-randomised studies**

| **Author** | **Country/region** | **Study Design** | **Selection** | **Comparability** | **Outcome** | **Total score** | **Result** |
| --- | --- | --- | --- | --- | --- | --- | --- |
| Bailey et al. (2022) ^33^ | Kenya | Cross-sectional | *** | ** | ** | 7 | Good |
| Beckham et al. (2023) ^34^ | US | Cross-sectional | *** | ** | ** | 7 | Good |
| Beymer et al. (2018) ^26^ | US | Cohort study | *** | ** | ** | 7 | Good |
| Biello et al. (2023) ^49^ | US | Cross-sectional | *** | ** | ** | 7 | Good |
| Calabrese et al. (2019) ^56^ | US | Cross-sectional | *** | ** | ** | 7 | Good |
| Chan et al. (2022) ^35^ | Australia | Cross-sectional | *** | ** | ** | 7 | Good |
| Dubov er al. (2018) ^64^ | Ukraine | Cross-sectional | *** | ** | ** | 7 | Good |
| Dubov er al. (2019) ^45^ | US | Cross-sectional | ** | ** | ** | 6 | Fair |
| Ellison et al. (2019) ^29^ | US | Cross-sectional | *** | ** | ** | 7 | Good |
| Fu et al. (2023) ^36^ | China | Cross-sectional | ** | ** | ** | 6 | Fair |
| Greene et al. (2017) ^58^ | US | Cross-sectional | *** | ** | *** | 8 | Good |
| Guo et al. (2023) ^37^ | China | Cross-sectional | ** | ** | ** | 6 | Good |
| Guttierrez et al. (2022) ^65^ | US | Cross-sectional | *** | ** | ** | 7 | Good |
| Hsu et al. (2023) ^38^ | Taiwan, China | Cross-sectional | *** | ** | ** | 7 | Good |
| Irie et al. (2022) ^53^ | US | Cross-sectional | *** | ** | *** | 8 | Good |
| John et al. (2018) ^27^ | US | Cross-sectional | ** | ** | ** | 6 | Fair |
| Luecke et al. (2016) ^52^ | Multicenter | Mixed-method | *** | ** | ** | 7 | Good |
| Mansergh et al. (2021) ^32^ | US | Cross-sectional | *** | ** | ** | 7 | Good |
| Martinez et al. (2022) ^39^ | Northeast Corridor region between Philadelphia and Trenton | Cross-sectional | *** | ** | ** | 7 | Good |
| Meyers et al. (2014) ^44^ | US | Cohort study | *** | ** | ** | 7 | Good |
| Meyers et al. (2016) ^50^ | US | Cross-sectional | *** | ** | ** | 7 | Good |
| Meyers et al. (2018) ^28^ | US | Cross-sectional | *** | ** | ** | 7 | Good |
| Meyers et al. (2018) ^28^ | China | Cross-sectional | *** | ** | ** | 7 | Good |
| Minnis et al. (2019) ^66^ | South Africa | Cross-sectional | *** | ** | ** | 7 | Good |
| Ogunbajo et al. (2022) ^40^ | Nigeria | Cross-sectional | *** | ** | ** | 7 | Good |
| Oldenburg et al. (2016) ^25^ | Vietnam | Cross-sectional | ** | ** | ** | 6 | Fair |
| Peng et al. (2019) ^30^ | China | Cross-sectional | *** | ** | ** | 7 | Good |
| Raccagni et al. (2023) ^42^ | Italy | Cross-sectional | *** | ** | ** | 7 | Good |
| Restar et al. (2023) ^57^ | Philippines | Cross-sectional | ** | ** | ** | 6 | Fair |
| Schoenberg et al. (2023) ^24^ | US | Cross-sectional | *** | ** | ** | 7 | Good |
| Slama et al. (2023) ^46^ | France | Cross-sectional | *** | ** | ** | 7 | Good |
| Stephenson et al. (2022) ^41^ | US | RCT | *** | ** | ** | 7 | Good |
| Tagliaferri et al. (2022) ^67^ | US | Cross-sectional | *** | ** | ** | 7 | Good |
| Tan et al. (2021) ^68^ | Canada | Cross-sectional | *** | ** | ** | 7 | Good |
| Tolley et al. (2019) ^55^ | Multicenter | Mixed-method | *** | ** | *** | 8 | Good |
| Tolley et al. (2020) ^69^ | US | Cross-sectional | ** | ** | ** | 6 | Fair |
| Torres et al. (2023) ^31^ | Multicenter | Cross-sectional | *** | ** | ** | 7 | Good |
| Valente et al. (2022) ^70^ | US | Cross-sectional | *** | ** | ** | 7 | Good |
| Wang et al. (2024) ^43^ | The Netherlands | Cross-sectional established within a cohort study | *** | ** | ** | 7 | Good |
| Wara et al. (2023) ^54^ | Africa | Observational cohort study / Observational extension cohort of a cluster randomized trial study | *** | ** | *** | 8 | Good |

The selection, comparability, and exposure of each study were broadly assessed. Studies with 3 or 4 stars in selection domain AND 1 or 2 stars in comparability domain AND 2 or 3 stars in outcome/exposure domain were considered of good quality; studies with 2 stars in selection domain AND 1 or 2 stars in comparability domain AND 2 or 3 stars in outcome/exposure domain were considered of fair quality; or were considered as poor quality.
